# Mechanistic models of Rift Valley fever virus transmission dynamics: A systematic review

**DOI:** 10.1101/2022.03.28.22272741

**Authors:** Hélène Cecilia, Alex Drouin, Raphaëlle Métras, Thomas Balenghien, Benoit Durand, Véronique Chevalier, Pauline Ezanno

**Author notes:** contributed equally (Hélène Cecilia, Alex Drouin).

## Abstract

Rift Valley fever (RVF) is a zoonotic arbovirosis which has been reported across Africa including the northernmost edge, South West Indian Ocean islands, and the Arabian Peninsula. The virus is responsible for high abortion rates and mortality in young ruminants, with economic impacts in affected countries. To this day, RVF epidemiological mechanisms are not fully understood, due to the multiplicity of implicated vertebrate hosts, vectors and ecosystems. In this context, mathematical models are useful tools to develop our understanding of complex systems, and mechanistic models are particularly suited to data-scarce settings. In this work, we performed a systematic review of mechanistic models studying RVF, to explore their diversity and their contribution to the understanding of this disease epidemiology. Researching Pubmed and Scopus databases (October 2021), we eventually selected 48 papers, which needed to provide a clear description of a mechanistic model with numerical application to RVF. We categorized models as theoretical, applied or grey, according to their will to represent a specific geographical context and their use of data to fulfill this intention. We discussed their contributions to the understanding of RVF epidemiology, and highlighted that theoretical and applied models can use different tools to meet common objectives. Through the examination of model features, we identified research questions left unexplored across scales, along with a substantial lack of justification when choosing a functional form for the force of infection. Overall, we showed a great diversity in RVF models, leading to substantial progress in our comprehension of epidemiological mechanisms. To go further, data gaps must be fulfilled, and modelers need to go the extra mile regarding transparency.

**Authors summary:** Rift Valley fever (RVF) affects humans and livestock across Africa, South West Indian Ocean islands, and in the Arabian Peninsula. This disease is one of the World Health Organization priorities, and is caused by a virus transmitted by Aedes and Culex spp. mosquitoes, but also directly from livestock to humans. Mathematical models have been used in the last 20 years to disentangle RVF virus transmission dynamics. These models can further our understanding of processes driving outbreaks, test the efficiency of control strategies, or even anticipate possible emergence. Provided with detailed datasets, models can tailor their conclusions to specific geographical contexts and aid in decision-making in the field. This review provides a general overview of mathematical models developed to study RVF virus transmission dynamics. We describe their main results and methodological choices, and identify hurdles to be lifted. To offer innovative animal and public health value, we recommend that future models focus on the relative contribution of host species to transmission, and the role of animal mobility.

## Introduction

Rift Valley fever (RVF) is a viral, vector-borne, zoonotic disease, first identified in Kenya in 1930 [1]. It has since then been reported across the African continent, in the South West Indian Ocean islands, and in the Arabian Peninsula. Rift Valley fever virus (RVFV) is mostly transmitted by *Aedes* and *Culex* spp. mosquitoes [2], some of which are present in Europe and North America [3–6]. In livestock, abortion storms and death can strongly impact the local economy [7,8]. Human infections arise mostly following contact with tissues of infected animals but can also be vector-mediated. The clinical spectrum in humans is broad, with a minority of deadly cases [9,10].

About 100 years after its first description, RVF outbreaks are still difficult to anticipate and contain, and the drivers of RVF endemicity are not clearly understood. The multiplicity of vertebrate host and mosquito species involved, the diversity of affected ecosystems, each with their own environmental dynamics, as well as the impact of human activities, make this complex system hard to disentangle [11]. The limited use of available vaccines [12], coupled with the overall social vulnerability of affected regions [13,14], are also major obstacles. The pastoralist tradition, which constitutes the main production system in African drylands [15], can induce delayed access to health care and hinder the traceability of animal mobility. This, in turn, impacts the quality and the availability of epidemiological data, which can be quite heterogeneous [16,17]. As a result, it is often difficult to generalize local findings, unless a mechanistic understanding of epidemiological processes is acquired.

Mathematical models are useful to project epidemiological scenarios, including control strategies, at large scales, be they temporal, spatial or demographic [18,19]. Powerful methods can now estimate the most likely drivers of observed outbreak patterns [20,21], or point out key processes needing further field or laboratory investigations [22]. Phenomenological models (used here to avoid the often-used but misleading term ‘statistical models’, [23] are effective to extract patterns from data. In contrast, mechanistic (or dynamical) models can adapt to data-scarce settings by exploring a complex system conceptually, in a hypothesis-driven fashion [24], e.g. to see what ranges of behavior can emerge from first principles, as is routinely done in ecology [25]. This flexibility gives rise to an interesting variability in the way epidemiological mechanistic models are designed and used, spanning a broad spectrum from highly theoretical to closely mimicking field situations [26].

Two existing reviews have focused on models developed to study RVF. The first one, by Métras et al. (2011) [27], was a narrative review presenting modeling tools used to measure or model RVF risk in animals. At that time, only three mechanistic models were available and included in the study. The second one, by Danzetta et al. (2016) [28], was a systematic review constrained to compartmental models which included 24 articles. The authors used RVF as a case study to present how the use of compartmental models can be helpful to investigate various aspects of vector-borne disease transmission. A complementary paper, by Reiner et al. (2013) [29], reviewed 40 years of mathematical models of mosquito-borne pathogen transmission, with a thorough and comprehensive reading grid. It did however only include three models on RVF.

To update the state of the art on mechanistic models of RVF virus transmission, we conducted a systematic review. Our main goal was to identify knowledge gaps left unaddressed in models, and therefore identify future research avenues. To achieve this, we categorized models in theoretical and applied models, and explored these categories throughout the paper to identify what they have in common and how they differ. First, we explored their inheritance connections and assessed whether these categories inspired each other. We then detailed their contribution to the understanding of RVF epidemiology. Lastly, we described the diversity of methodological choices and assumptions made in these models. In particular, we dedicated a whole section to present the different functional forms used by models for the force of infection. We detailed the underlying assumptions on host-vector interactions that these functional forms imply, as we noticed a lack of justification regarding this choice in reviewed papers, even though host-vector interactions represent a key factor in RVFV transmission.

## Material and Methods

### Search strategy

This review was conducted according to the Preferred Reporting Items for Systematic reviews and Meta-Analyses (PRISMA) guidelines [30,31]. The research was performed in Scopus and Pubmed databases on 12 October 2021. No restriction on publication date was considered. The following Boolean query was applied in both databases: (*rift* AND *valley* AND *fever*) AND (*mathematical* OR *epidem*\* OR *compartment*\* OR *sir* OR *seir* OR *metapopulation* OR *deterministic* OR *stochastic* OR *mechanistic* OR *dynamic*\*) AND (*model*\*)

This query was used in the “title, abstract and keywords”, and “title and abstract” fields for Scopus and PubMed, respectively.

### Inclusion and exclusion criteria

After removal of duplicates, studies were included in three steps: title screening, abstract screening and full text reading. In the first and second step, records were selected if they appeared to present a RVF model using a mechanistic approach for at least one part of the model. Exclusion criteria were: irrelevant topic, reviews, cases report, serological studies and statistical studies. Records selected in the first and second step went to a full text screening of the corresponding report, with the first set of exclusion criteria, with addition of: non-mechanistic models, models dealing with mosquitoes only, incomplete model description and theoretical paper without any RVF numerical application. Discussion among authors occurred in case of doubt to reach a consensus on final inclusions.

### Screening

We designed a reading grid (Supplementary Information Text S.1), partially inspired by the one used in Reiner et al. (2013) [29], to collect information from the studies. The context of the study (e.g. location, presence of data), model components (e.g. host and vector species, infection states) and assumptions (e.g. vertical transmission in vectors), type of outputs (e.g.*R*_O_, parameter estimations, sensitivity analysis), and main results were all recorded. Two authors took charge of the systematic reading. To cross-validate the use of the grid, three studies were read by both authors and specific topics were regularly discussed to make sure a consensus was reached.

### Model typology and inheritance connections

We defined three model categories: theoretical, applied, and grey models. Theoretical models do not use any data and are not intended to represent any specific geographical location. Applied models represent a specific geographical context and use relevant data to tailor model development to their case study or to validate model outputs. Such data can be of several types, as environmental or demographic data, and not necessarily epidemiological in the sense of seroprevalence or case reports. Grey models are those which do not fit into these well-defined categories, either because authors do not use data but demonstrate a strong will to adapt their model to a specific geographical or epidemiological context, or because despite the use of data, the model developed is still very conceptual and insights remain distant from field preoccupations. We recorded inheritance connections between studies: if a model explicitly stated being adapted from another model, we defined the latter as a parent model.

## Results and discussion

### Study selection

A total of 372 records were identified from the two databases. After removal of duplicates, 248 records were screened at the title level, 146 at the abstract level and 69 reports were fully read. Eventually, 49 studies were selected for the present review (Figure 1). Among those, 26 were not present in the review by Danzetta et al. (2016) [28].

**Figure 1.**
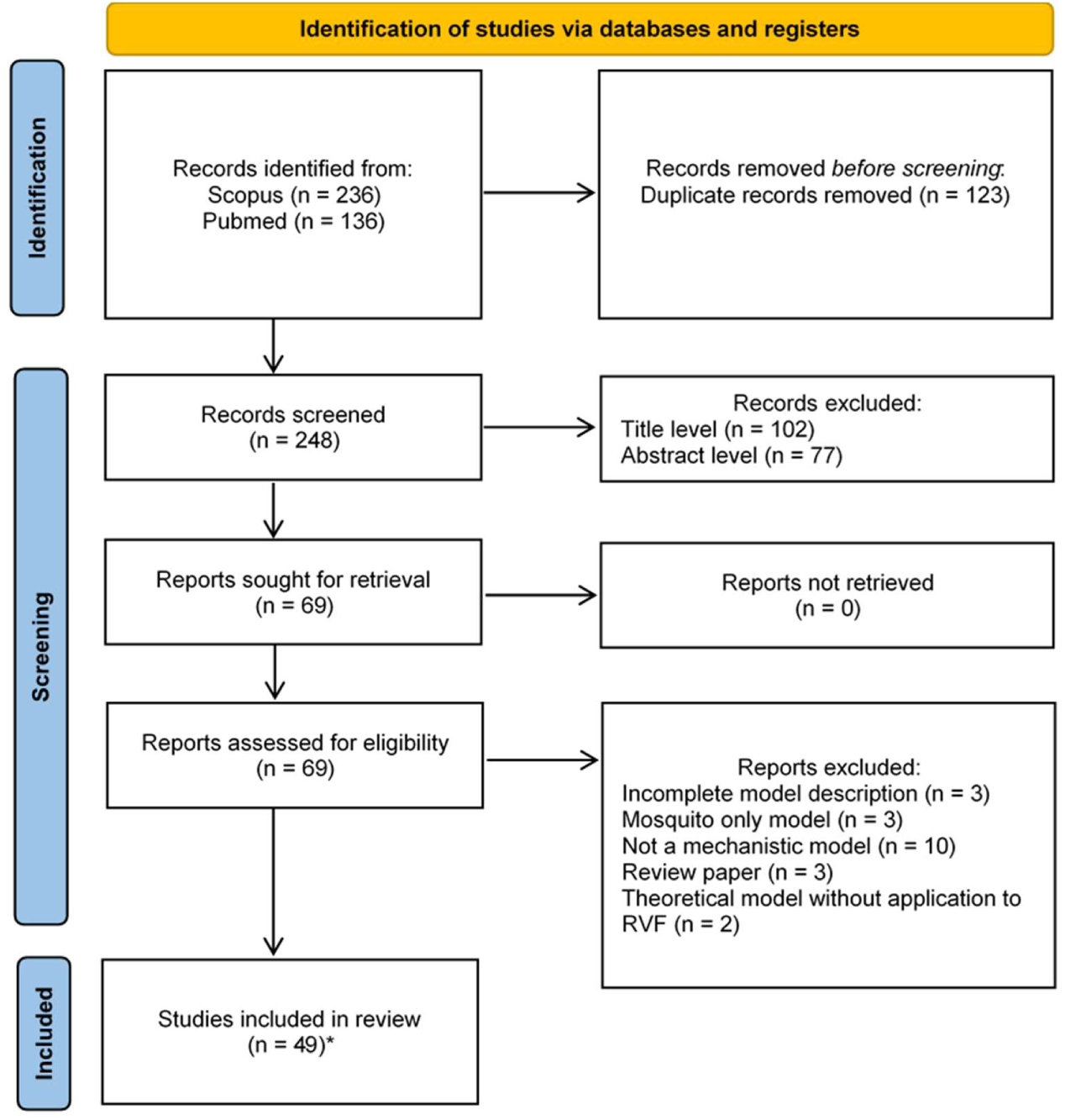
PRISMA flow diagram representing the selection process Record: title and/or abstract of a report indexed in a database Report: document supplying information about a study Study: An experiment, corresponding here to models [30] * One report included two studies

### Model typology and inheritance connections

We identified 18 applied models (37%), 18 theoretical models (37%), and 13 grey models (26%, Table 1). Twenty models (41%) declared a parent model within the list of presently reviewed studies, for a total of 26 models in the inspirational network (Figure 2). In 14 cases (70%), a model and its parent shared at least one author. In 13 cases, a model and its parent belonged to the same category (5 applied, 1 grey, 7 theoretical). The model by Gaff et al., (2007) [32] is a clear example of a model laying the groundwork for future model developments. It was first modified to explore several control strategies in Gaff et al. (2011) [33] (theoretical). Adongo et al. (2013) [34] (theoretical) then elaborated on Gaff et al. (2011) [33] to explore sophisticated vaccination schemes. Besides, Gaff et al., (2007) [32] model was spatialized in Niu et al. (2012) [35] (theoretical). In other cases, theoretical and grey studies provided a basis for the construction of more applied models in further work. One grey model [36] was the parent of an applied model [37]. In 4 cases, a theoretical model ([32] twice, [38], [39]) was a parent of a grey model ([40], [41], [42] and [36] respectively). Lastly, Gaff et al., (2007) [32], a theoretical model, was the parent of 2 applied models [43,44].

**Table 1:** Main characteristics of mechanistic models on Rift Valley fever virus transmission dynamics included in the review.

| Study | Model category | Primary objective | Main output | Deterministic or stochastic ? | Geographical zone | Scale | Number vertebrate hosts taxa | Taxa | Host health states | Number vector taxa | Taxa | Vector health states | FOI | Compartmental or ABM ? | Code access |
| --- | --- | --- | --- | --- | --- | --- | --- | --- | --- | --- | --- | --- | --- | --- | --- |
| Beechler et al. (2015) [37] | Applied | Understand | Scenario comparison | Deterministic | South Africa | Local | 1 | Buffalo | SIR | 1 | *Aedes* (assumed) | SEI | Hybrid1 | Compartmental | No |
| Bicout and Sabatier (2004) [54] | Applied | Understand | Scenario comparison | Deterministic with at least one stochastic process | Senegal | Local | 1 | Livestock | IR (other states not described) | 2 | *Aedes*, *Culex* | Not explicit | FR | Compartmental | No |
| Scoglio et al. (2016) [55] | Applied | Understand | Scenario comparison | Stochastic | United States of America | Sub-national | 1 | Cattle | SEIR | 0 |  |  | NA | ABM | Yes |
| Sekamatte et al. (2019) [56] | Applied | Understand | Scenario comparison | Stochastic | Uganda | Sub-national | 1 | Cattle | SEIR | 0 |  |  | NA | ABM | No |
| Leedale et al. (2016) [51] | Applied | Understand | Risk map | Deterministic | Kenya, Tanzania | International | 1 | Livestock | SEIR | 2 | *Aedes*, *Culex* | SEI | FR | Compartmental | No |
| Cecilia et al. (2020) [57] | Applied | Understand | Risk map | Deterministic | Senegal | Sub-national | 2 | Cattle, Small ruminants | SEIR | 2 | *Aedes*, *Culex* | SEI | FR | Compartmental | Yes |

Table 1 – Continued
| Study | Model category | Primary objective | Main output | Deterministic or stochastic ? | Geographical zone | Scale | Number vertebrate hosts taxa | Taxa | Host health states | Number vector taxa | Taxa | Vector health states | FOI | Compartmental or ABM ? | Code access |
| --- | --- | --- | --- | --- | --- | --- | --- | --- | --- | --- | --- | --- | --- | --- | --- |
| Xue et al. (2013) [46] | Applied | Understand | Risk map | Deterministic with at least one stochastic process | United States of America | Sub-national | 2 | Cattle, Human | SEIR | 2 | *Aedes*, *Culex* | SEI | FI | Compartmental | No |
| Xue et al. (2012) [44] | Applied | Understand | Parameter estimation | Deterministic | South Africa | Sub-national | 2 | Sheep, Human | SEIR | 2 | *Aedes*, *Culex* | SEI | FI | Compartmental | No |
| Nicolas et al. (2014) [59] | Applied | Understand | Parameter estimation | Deterministic | Madagascar | Sub-national | 1 | Cattle | SEIR | 1 | Not precised | SEI | FR | Compartmental | No (upon request) |
| Métras et al. (2017) [48] | Applied | Understand | Parameter estimation | Deterministic | Mayotte | Sub-national | 1 | Livestock | SEIR | 0 |  |  | NA | Compartmental | No |
| Métras et al. (2020) [47] | Applied | Understand | Parameter estimation | Deterministic | Mayotte | Sub-national | 2 | Livestock, Human | SEIRVsVp | 0 |  |  | NA | Compartmental | No |
| Tennant et al. (2021) [60] | Applied | Understand | Parameter estimation | Deterministic | Comoros archipelago | International | 1 | Livestock | SEIR | 0 |  |  | NA | Compartmental | Yes |
| Durand et al. (2020) [61] | Applied | Understand | Parameter estimation | Deterministic with at least one stochastic process | Senegal | Local | 2 | Cattle, Small ruminants | SIR | 2 | *Aedes*, *Culex* | SEI | FR | Compartmental | No |
| Barker et al. (2013) [43] | Applied | Anticipate | Risk map | Deterministic | United States of America | Sub-national | 2 | Cattle, Birds | SEIR | 2 | *Aedes*, *Culex* | SEI | FI | Compartmental | No |
| Fischer et al. (2013) [62] | Applied | Anticipate | Risk map | Deterministic | Netherlands | National | 2 | Cattle, Small ruminants | SEIR | 2 | *Aedes*, *Culex* | SEI | FR | Compartmental | No |

Table 1 – Continued
| Study | Model category | Primary objective | Main output | Deterministic or stochastic ? | Geographical zone | Scale | Number vertebrate hosts taxa | Taxa | Host health states | Number vector taxa | Taxa | Vector health states | FOI | Compartmental or ABM ? | Code access |
| --- | --- | --- | --- | --- | --- | --- | --- | --- | --- | --- | --- | --- | --- | --- | --- |
| EFSA AHAW Panel et al. (2020 – Model 1) [49] | Applied | Control | Scenario comparison | Stochastic | Mayotte | Sub-national | 1 | Livestock | SEIRV | 1 | *Culex* | SEI | NA | Compartmental | No |
| EFSA AHAW Panel et al. (2020 – Model 2) [49] | Applied | Control | Risk map | Stochastic | Netherlands | National | 1 | Livestock | SIR | 0 |  |  | FR | Compartmental | No |
| Gao et al. (2013) [53] | Grey | Understand | Scenario comparison | Deterministic | Sudan, Egypt | International | 1 | Livestock | SIR | 1 | Not precised | SI | MA | Compartmental | No |
| Manore and Beechler (2015) [36] | Grey | Understand | Scenario comparison | Deterministic | South Africa | Local | 1 | Buffalo | SIR | 1 | *Aedes* | SEI | Hybrid1 | Compartmental | No |
| Xiao et al. (2015) [52] | Grey | Understand | Scenario comparison | Deterministic | Sudan, Egypt | International | 1 | Livestock | SEIR | 1 | *Culex* | SEI | MA | Compartmental | No |
| Lo Iacono et al. (2018) [63] | Grey | Understand | Scenario comparison | Deterministic | Kenya | National | 1 | Livestock | SEIR | 2 | *Aedes, Culex* | SEI | Hybrid3 | Compartmental | No |
| Sumaye et al. (2019) [64] | Grey | Understand | Scenario comparison | Deterministic | Tanzania | Sub-national | 2 | Cattle, Human | SEIR | 4 | *Aedes, Culex* | SI | Hybrid1 | Compartmental | Yes |

Table 1 – Continued
| Study | Model category | Primary objective | Main output | Deterministic or stochastic ? | Geographical zone | Scale | Number vertebrate hosts taxa | Taxa | Host health states | Number vector taxa | Taxa | Vector health states | FOI | Compartmental or ABM ? | Code access |
| --- | --- | --- | --- | --- | --- | --- | --- | --- | --- | --- | --- | --- | --- | --- | --- |
| McMahon et al. (2014) [65] | Grey | Understand | Scenario comparison | Deterministic with at least one stochastic process | East Africa | International | 3 | Cattle, Wildlife, Human | SEIRAV | 2 | *Aedes*, *Culex* | SEI | Hybrid2 | Compartmental | No |
| Gil et al. (2016) [66] | Grey | Understand | Scenario comparison | Both tested | Egypt | National | 1 | Livestock | SIR | 1 | *Culex* | SI | MA | Both tested | No |
| Tuncer et al. (2016) [67] | Grey | Understand | Parameter estimation | Deterministic | Kenya | Sub-national | 2 | Livestock, Human | SI-R for livestock only | 1 | Not precised | SI | MA* | Compartmental | No |
| Cavalerie et al. (2015) [41] | Grey | Understand | Parameter estimation | Stochastic | Mayotte | Sub-national | 1 | Livestock | SEIR | 1 | Mean from several species | SEI | FR | Compartmental | Yes |
| Mpeshe, Luboobi and Nkansah-Gyekye (2014) [42] | Grey | Understand | Sensitivity analysis | Deterministic | Tanzania | Sub-national | 2 | Livestock, Human | SEIR | 2 | *Aedes*, *Culex* | SEI | FI | Compartmental | No |
| Pedro, Abelman and Tonnang (2016) [68] | Grey | Understand | Mathematical properties | Stochastic | East Africa, South Africa |  | 1 | Livestock | SIR | 1 | *Aedes* | SI | Hybrid1 | ABM | No |
| Miron et al. (2016) [40] | Grey | Anticipate | Sensitivity analysis | Deterministic | North America | Local | 2 | Livestock, Human | SEIR | 1 | *Aedes* | SEI | MA | Compartmental | No |
| Gachohi et al. (2016) [69] | Grey | Control | Scenario comparison | Deterministic | Kenya | Local | 2 | Cattle, Small ruminants | SEIR | 2 | *Aedes*, *Culex* | SEI | FR | Compartmental | Yes |

Table 1 – Continued
| Study | Model category | Primary objective | Main output | Deterministic or stochastic ? | Geographical zone | Scale | Number vertebrate hosts taxa | Taxa | Host health states | Number vector taxa | Taxa | Vector health states | FOI | Compartmental or ABM ? | Code access |
| --- | --- | --- | --- | --- | --- | --- | --- | --- | --- | --- | --- | --- | --- | --- | --- |
| Niu et al. (2012) [35] | Theoretical | Understand | Scenario comparison | Deterministic |  |  | 1 | Livestock | SEIR | 2 | *Aedes, Culex* | SEI | FI | Compartmental | No |
| Chamchod et al. (2014) [70] | Theoretical | Understand | Scenario comparison | Deterministic |  |  | 1 | Livestock | SIR | 1 | Not precised | SI | FR | Compartmental | No |
| Pedro (2018) [71] | Theoretical | Understand | Scenario comparison | Deterministic |  |  | 1 | Livestock | SIR | 1 | *Aedes* | SI | FR | Compartmental | No |
| Wen, Teng and Liu (2019) [72] | Theoretical | Understand | Scenario comparison | Deterministic |  |  | 1 | Livestock | SEIR | 1 | Not precised | SEI | MA | Compartmental | No |
| Python Ndekou Tandong et al. (2020) [58] | Theoretical | Understand | Scenario comparison | Deterministic |  | Local | 1 | Animals | SEIR | 2 | *Aedes, Culex* | SEI | FI | Hybrid<sup>1</sup> | No |
| Mpeshe (2021) [73] | Theoretical | Understand | Scenario comparison | Deterministic |  |  | 1 | Human | SEIR | 1 | Not precised | SEI | FI | Compartmental | No |
| Xue and Scoglio (2015) [74] | Theoretical | Understand | Scenario comparison | Deterministic with at least one stochastic process |  |  | 1 | Livestock | SEIR | 1 | Not precised | SEI | FR | Compartmental | No |
| Gaff, Hartley and Leahy (2007) [32] | Theoretical | Understand | Sensitivity analysis | Deterministic |  |  | 1 | Livestock | SEIR | 2 | *Aedes, Culex* | SEI | FI | Compartmental | No |
| Mpeshe, Haario and Tchuente (2011) [38] | Theoretical | Understand | Sensitivity analysis | Deterministic |  |  | 2 | Livestock, Human | SEIR | 1 | Not precised | SEI | FI | Compartmental | No |
| Chitnis, Hyman and Manore (2013) [39] | Theoretical | Understand | Sensitivity analysis | Deterministic |  |  | 1 | Cattle | SIR | 1 | *Aedes* | SEI | Hybrid1 | Compartmental | No |
| Xue and Scoglio (2013) [75] | Theoretical | Understand | Sensitivity analysis | Deterministic |  |  | 2 | Livestock, Human | SEIR | 2 | *Aedes*, *Culex* | SEI | FI | Compartmental | No |
| Pedro, Tonnang and Abelman (2016) [45] | Theoretical | Understand | Sensitivity analysis | Deterministic |  |  | 1 | Livestock | SIRA | 2 | *Aedes*, *Culex* | SEI | Hybrid1 | Compartmental | No |
| Pedro, Abelman sand Tonnang (2017) [76] | Theoretical | Understand | Sensitivity analysis | Deterministic |  |  | 1 | Livestock | SIR | 3 | *Aedes*, *Culex*, *Hyalomma* ticks | SE (Mosquitoes only) I | Hybrid1 | Compartmental | No |
| Pedro et al. (2014) [77] | Theoretical | Understand | Mathematical properties | Deterministic |  |  | 1 | Livestock | SIRA | 2 | *Aedes*, *Culex* | SEI | Hybrid1 | Compartmental | No |
| Gaff et al. (2011) [33] | Theoretical | Control | Scenario comparison | Deterministic |  |  | 1 | Cattle | SEIRV | 2 | *Aedes*, *Culex* | SEI | FI | Compartmental | No |
| Adongo et al. (2013) [34] | Theoretical | Control | Scenario comparison | Deterministic |  |  | 1 | Livestock | SEIR | 2 | *Aedes*, *Culex* | SEI | FI | Compartmental | No |

Table 1 - Continued
| Study | Model category | Primary objective | Main output | Deterministic or stochastic ? | Geographical zone | Scale | Number vertebrate hosts taxa | Taxa | Host health states | Number vector taxa | Taxa | Vector health states | FOI | Compartmental or ABM ? | Code access |
| --- | --- | --- | --- | --- | --- | --- | --- | --- | --- | --- | --- | --- | --- | --- | --- |
| Chamchod et al. (2016) [78] | Theoretical | Control | Scenario comparison | Deterministic |  |  | 1 | Livestock | SIRV1V2 | 1 | Not precised | SI | MA | Compartmental | No |
| Yang and Nie (2016) [79] | Theoretical | Control | Scenario comparison | Deterministic |  |  | 1 | Livestock | SIR | 1 | Not precised | SI | MA | Compartmental | No |

**Figure 2.**
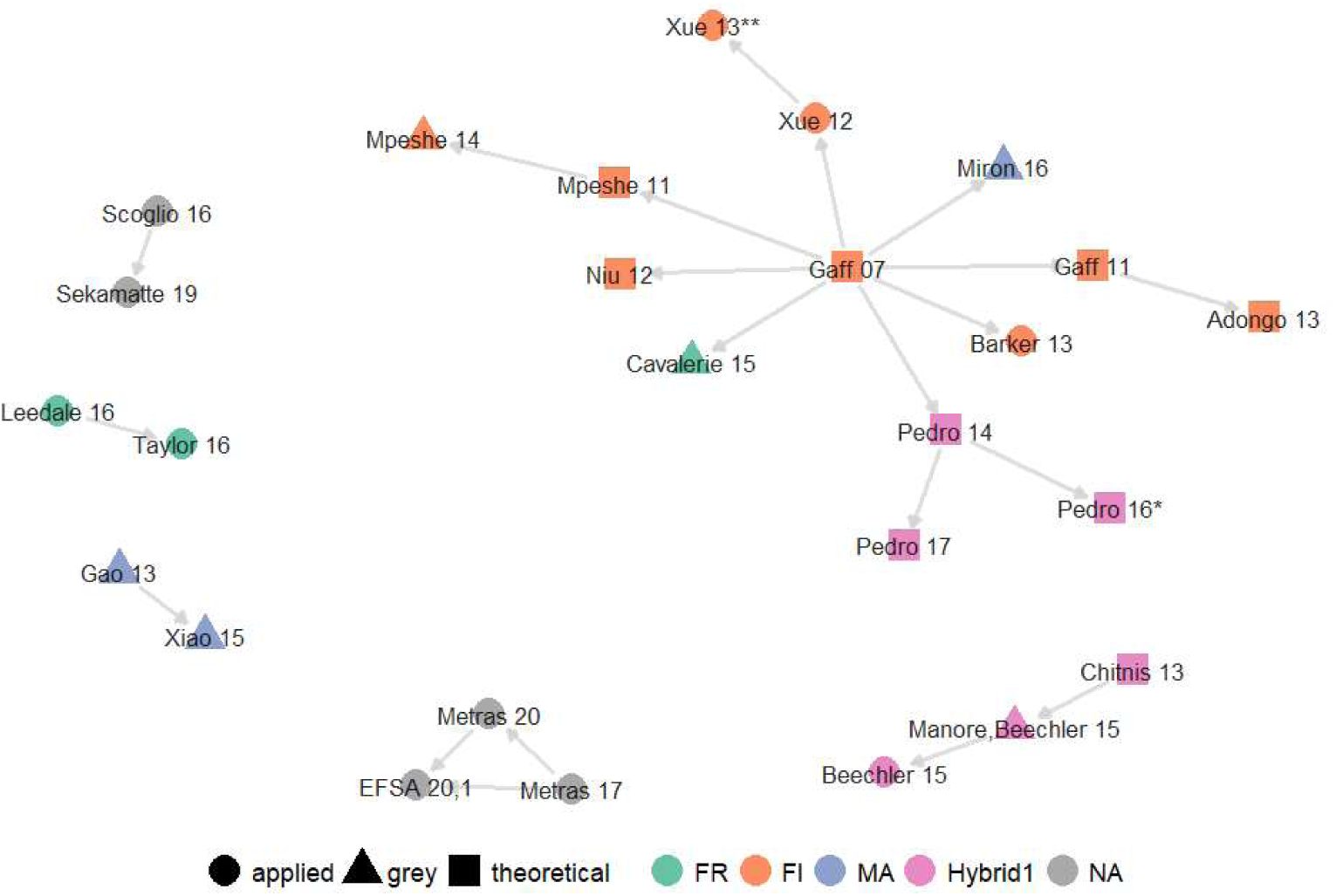
Inspirational network of models. Nodes are labeled with the reference of the associated studies (year abbreviated), shaped by model category, and colored by the functional form of the force of infection (see section on Force of infection). An edge between two nodes represents a model declaring the other as its parent model, as defined in the main text. Twenty-three models are not shown in this plot as they did not declare a parent model within the list of presently reviewed studies. *[45] ** [46]

Changes in model features can also give an overview of the continuity between a model and its parent. Métras et al, (2020) [47] added a human compartment to the model of Métras et al. (2017) [48] and ran the parameter estimation algorithm on a new outbreak dataset. One of the two models described in EFSA AHAW Panel et al., (2020)[49] then made a stochastic model based on Métras et al. (2017, 2020) [47,48]. Taylor et al., (2016) [50] used the model by Leedale et al. (2016) [51] to explore a new research question (anticipate the effect of climate change) in an expanded region (East African Community compared to Kenya and Tanzania). Xiao et al. (2015) [52] modified the model by Gao et al. (2013) [53] to include seasonality through time-varying parameters.

### Contribution to the understanding of RVF epidemiology

#### Objective of the modeling study

To broadly describe the contribution of models to the study of RVF epidemiology, three main scientific objectives were identified (Table 1): exploring epidemiological mechanisms (‘understand’, n = 38), examining consequences of hypothetical outbreaks (‘anticipate’, n = 4), and assessing control strategies(‘control’, n = 7). In the present section, we focus on key features identified per objective.

The most common primary scientific objective of models was to understand epidemiological processes, in all model categories (from 72% of applied models to 79% of grey models, Table 1, Figure 3). Although in 11 cases, those models also aimed to anticipate or control outbreaks in a secondary part [32,47,48,55–58,60,65,68,70]. In addition, in 30% of cases, model development in itself seemed to be a leading objective of the study. In such cases, contributing to RVF epidemiology was as important as contributing methodologically to RVF mechanistic modeling, by including for the first time a given compartment, parameter, or by developing a method to integrate data.

**Figure 3.**
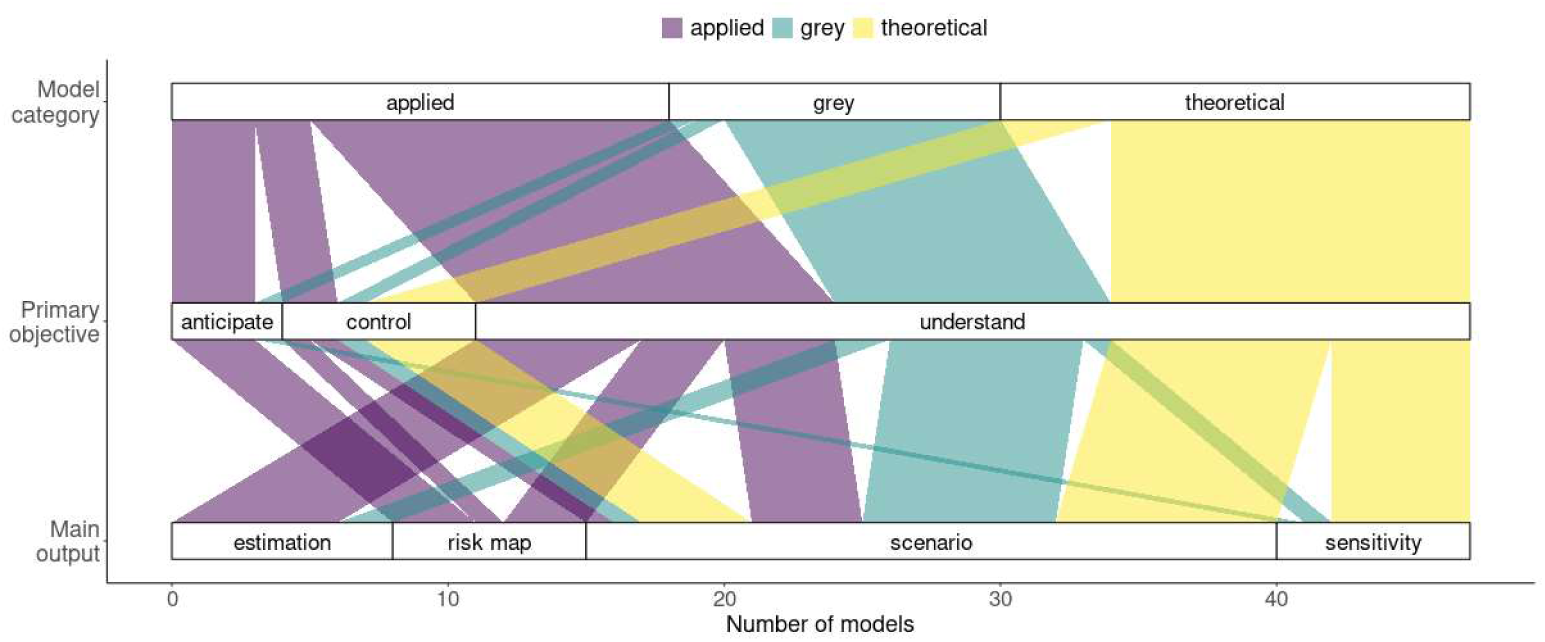
Association between the model category, the primary objective of the study, and the main type of output chosen to illustrate the results. This figure excludes two models for which the main output consisted of a deep analysis of the mathematical properties of the system (Table 1).

An interesting trend is the evolution of the objectives of modeling papers over the years, which increasingly include the control and anticipation of RVF outbreaks (15/26 studies in 2016-present, 7/23 in 2004-2015). Research on RVF, through mathematical modeling and other methods, has deeply enhanced our understanding of underlying epidemiological mechanisms, which now allows models to focus more on operational aspects. However, some papers did not formulate a precise research question and consequently did not tailor their model to a specific set of hypotheses or scenarios to test. Theoretical models have helped to broadly explore the pathosystem behavior when dealing with a lot of uncertainty, but such papers often lack clarity. A difficulty for theoretical papers is to convey how mathematical analysis can be helpful to field practitioners down the line. Regarding applied and grey models, their specificity often relied on the geographical application and the dataset they used, rather than on a focused research question.

#### Main outputs

The main output of a model, holding the key message of the studies, could pertain to one of five categories (Table 1) : i) parameter estimation (n = 8), ii) risk maps (n = 7), iii) comparison of scenarios, defined as a small set of simulations with specific parameters varied (or processes turned off) across a small set of values (n = 25), and iv) sensitivity analysis, where a large subset (if not all) of parameters are varied across a large set of values (e.g. using sampling design to generate them), usually to produce an index quantifying the impact of each parameter on selected model outputs (n = 7). In 2 cases, the main results relied on a deep analysis of the mathematical properties of the system (e.g. van Kampen system-size expansion [68], Lyapunov exponent, Poincaré map [77]). A given paper could have produced several of these outputs but we tried to identify, with an inevitable part of subjectivity, the one standing out as the main output.

The main model output varied according to the model category and their primary objective (Figure 3). Scenario comparison was the only main output used by all model categories (Figure 3). Indeed, this type of analysis is flexible and can focus on a specific hypothesis and its impact on the system’s behavior. Risk maps were only produced as a main output by applied models, and was the most common output for models with the aim to anticipate (Figure 3). Sensitivity analyses were mostly used by theoretical models as a main output (5/7), and never as such by applied models (Figure 3, 3/10 by grey models). Parameter estimation was mostly performed by applied models (6/8), and not at all by theoretical models (Figure 3, 2/8 by grey models). By nature, sensitivity analyses and parameter estimation are primarily done to understand the system better. Here, we highlight that theoretical and applied models can use different tools to contribute to a common objective. In 3 cases, parameter estimations were used further in the same model to help anticipate [48] or control [47,60] outbreaks, as a secondary objective. Fifty-five percent of models provided an estimation of a type of reproduction number (e.g., the basic reproduction number *R*_O_, the effective reproduction number *R*_e_, the seasonal reproduction number *R*_st_ (phenomenological relationship estimated between environmental parameters and transmission rate) or the Floquet ratio *R*_T_ (the expected number of cases caused by a primary case after one complete cycle of seasons [80]), most of which were obtained analytically (25/27).

#### Key questions

Mechanistic models can help to gauge the importance of hardly observable epidemiological processes, such as vertical transmission in vectors. This transmission route was included in around 50% of models, all having ‘understand’ as a main objective of the study, which seems representative of current knowledge on the importance of this process in the field. Indeed, evidence is limited regarding its potential role in the interepidemic maintenance of the virus [81]. Five models centered their research question on the quantification of this mechanism, in all categories (2 theoretical, 2 grey, 1 applied). Chitnis et al. (2013) [39] (theoretical) showed that while the vertical transmission rate does not impact *R*_0_, it can contribute significantly to inter-epidemic persistence. Pedro et al. (2016) [45] (theoretical) estimated a linear significant effect of vertical transmission on R_0_ and vector eradication effort, although this effect became substantial only when vertical transmission rate was above 10%. Manore & Beechler (2015) [36] (grey) focused on inter-epidemic activity in Kruger National Park (South Africa) and estimated that realistic vertical transmission rates should be combined with the presence of alternate hosts to allow RVF persistence. Lo Iacono et al. (2018) [63] (grey) showed that transovarial transmission of RVF virus in *Aedes* spp. was not a prerequisite for RVF persistence over time in Kenya. Durand et al., (2020) [61] (applied) concluded that vertical transmission could not be ruled out but nomadic herd movements were sufficient to explain the endemic circulation of RVF virus in Senegal.

The importance of animal movements in RVF spread and persistence is another key question explored by included studies. Theoretical models shows that local and distance spread of the virus are positively correlated to animal movement speed and flow size [52,58], but complex relationships exist in case of heterogeneous movements and livestock death rates [75]. Spatial spread can also be limited by physical barriers to livestock migration [35]. Role of animal movements in RVF spread is also highlighted by applied models, especially with a low transmission probability [56] or in a low vectorial capacity [55] context. Métras et al. (2017) [48] suggested that import of infected livestock in 2007 was a major driver of RVF emergence in Mayotte in 2008-2010 and Gao et al. (2013) [53] that a transport of only few infectious animals from Sudan to Egypt are sufficient to start an outbreak. In Grande Comore, the RVF virus seems to be able to circulate even in absence of new introduction [60].

Original research questions stood out from the rest. Beechler et al. (2015) [37] studied the impact of co-infections with another pathogen (Bovine tuberculosis, BTB). Their data highlighted that RVF virus infection was twice as likely in BTB+ than BTB-individuals. Once this effect was incorporated in a model, an increase in BTB prevalence nonlinearly affected RVF outbreak size in both BTB-infected and BTB-free populations. Pedro et al. (2017) [76] looked at the possible role of ticks as vectors in addition to mosquitoes. They concluded that if ticks were capable of carrying and transmitting RVFV, this sensibly changed the transmission dynamics. Specifically, the size of outbreaks was increased, with a higher peak, reached faster, and the outbreak duration was reduced, compared to a situation with only mosquito vectors. Tuncer et al. (2016) [67] developed an immuno-epidemiological model in which pathogen load impacted transmission rate, and focused on the identifiability of parameters (i.e. the uniqueness of parameter values able to reproduce a given model trajectory) rather than the epidemiological impact of such a hypothesis.

A single model [50] has looked at the possible effect of climate change on RVF risk, in Eastern Africa. This likely does not reflect a lack of interest for this issue, but could rather indicate that mechanistic modeling is not the preferred method to study such trends, compared to phenomenological (i.e. statistical) models [82–85]. In their review, Métras et al. (2011) [27] had highlighted the widespread use of phenomenological models to assess RVF risk across spatio-temporal scales. Phenomenological models can play a key role in selecting relevant processes to include or characterize suitable habitats, by highlighting significant correlations in complex datasets [86–88]. Such phenomenological models can then be nested into mechanistic models for specific processes (e.g., temperature-dependency, density-dependency). Mechanistic and phenomenological approaches can be seen as complementary ways to build a comprehensive view of vector-borne and zoonotic pathosystems [89]. Still, how to prioritize research on livestock and human health in the context of climate change is up to debate [90,91].

#### Control measures

Currently, vaccination against RVF is only available for livestock, using live attenuated virus or inactivated virus vaccines, but with limitations in their use [92]. Ten models reflected on possible vaccination strategies (Table A in S1 Supporting Information), in all categories (3 applied, 5 theoretical, 2 grey). Main objectives of all of these studies was to ‘control’, except for Métras et al. (2020) [47] for which it was a secondary objective. Such strategies were shaped by parameters such as the time to build-up immunity, vaccine efficacy, coverage, and regimen (Table A in S1). Most models confirmed quantitatively the intuitive need for vaccination to happen before outbreaks or quickly after the first cases are detected, to have a significant impact (Table A in S1). EFSA AHAW Panel et al., (2020 - Model 1), Gachohi et al. (2016) and Métras et al. (2020) [47,49 - Model 1,69] incorporated constraints on the number of individuals vaccinated per day, so that a given coverage is reached at a realistic pace. Regarding the choice of hosts to vaccinate, Gachohi et al. (2106) [69] highlighted that while small ruminants needed a smaller coverage than cattle to achieve a given reduction in incidence, the vaccination of cattle provided the benefit of protecting both host populations. This important role of cattle in RVFV transmission is due to a higher vector to host ratio and a larger body surface area, attracting more mosquitoes. Métras et al. (2020) [47] estimated that, in the context of Mayotte island, vaccination of livestock was the most efficient strategy to limit human cases. It required fewer doses than human vaccination to achieve a similar reduction in cases, assuming a highly immunogenic, single dose, and safe vaccine was available in both populations. This model took into account human exposure to livestock in their risk of infection. Adongo et al. (2013) [34] showed that optimal strategies differed depending on whether one prioritized the minimization of costs (doses) or of infections, with no clear take-home message for policy makers. Chamchod et al. (2016) [78] explored differences between the use of live and killed vaccines, estimating that even with *R*_O_ < 1, RVF was likely to become endemic when live vaccines were implemented.

Vector control methods, using adulticides or larvicides are expensive and difficult to implement, due to the diversity of potential vector species and of breeding sites to treat [9,12]. These mitigations methods have been tested in a few models, with ambiguous results. Miron et al. (2016) [40] concluded that reducing mosquito lifetime under 8.67 days would reduce R0 below one. In some other cases, negative effects of vector control arose. If all compared strategies were efficient to reduce the number of cases in a context of high virus transmission, using larvicides in a low transmission context led to an increased incidence [33]. In Mayotte, mosquito abundance had to be decreased by more than 40% to reduce RVF incidence and epidemic length, and an increased duration of epidemics was observed with lower levels of control. In the same model, vector control showed efficiency when coupled with culling strategy [49 - Model 1].

Few models considered movement restriction as a control method. A reduction of movements led to a decrease in disease spatial spread [55] and in incidence [65], and can help to eradicate the disease [58]. In Uganda, Sekamatte et al. (2019) [56] concluded that during periods of low mosquito abundance, movement restrictions led to a significant reduction in incidence. Movement restrictions had little impact in case of high vector abundance if used alone, and should therefore be combined with mosquito control. However in some cases mitigating measures could have unexpected consequences. In Comoros, movement restriction between Grande Comore and other islands of the archipelago led to a more severe and delayed outbreak, and within-island controls appeared to be more effective [60].

In addition, testing and culling infected animals has been compared to other mitigations methods by three studies. This appeared to be one of the best strategies when conducted during 28 days after the detection of an outbreak in the theoretical model by Gaff et al. (2011) [33]. In the Netherlands, stamping out in a 20km radius around an outbreak was the most effective strategy compared to vaccination or other culling strategies [49 - Model 2]. Nevertheless in Mayotte an effective strategy seemed hard to implement due to the high level of animal testing and culling needed to be efficient [49 - Model 1].

Overall, modeling studies often (6 applied, 6 theoretical, 3 grey) incorporate control-like scenarios, but the applicability of such simulations can be improved. Few models tried to assess RVF mitigation strategies in real endemic settings, with 6 studies in areas with history of RVF circulation and only two with ‘prevent’ as a primary objective. Vaccination (n=10) and vector control (n=5) were the main strategies considered by models, although they currently present major on-field limitations (Nielsen et al. 2020). In addition, simulated vector control scenarios are often simplistic, consisting of a variation of one parameter homogeneously, and only one model distinguished between the use of larvicides and adulticides (Gaff et al. 2011). Finally, only five models considered movement restrictions as a mitigation strategy, which has been highlighted as a key determinant of RVF spread and persistence in some epidemiological contexts (Durand et al., 2020). Future efforts should focus on incorporating field constraints into their scenarios, while keeping in mind the transboundary nature of Rift Valley fever virus transmission.

### Model features

#### Geographical context

Locations of applied and grey models are mapped in Figure 4A. The scale of applied and grey models varied from local to international (Figure 4B). The sub-national scale was the most prevalent in both applied (10/18) and grey models (4/13) (Figure 4B). Regarding zones with known presence of RVF, several countries reporting numerous outbreaks in the last 15 years [12] have had at least one specific model developed. Besides, the Netherlands and the US, both RVF-free, were also used as case studies for several models.

**Figure 4.**
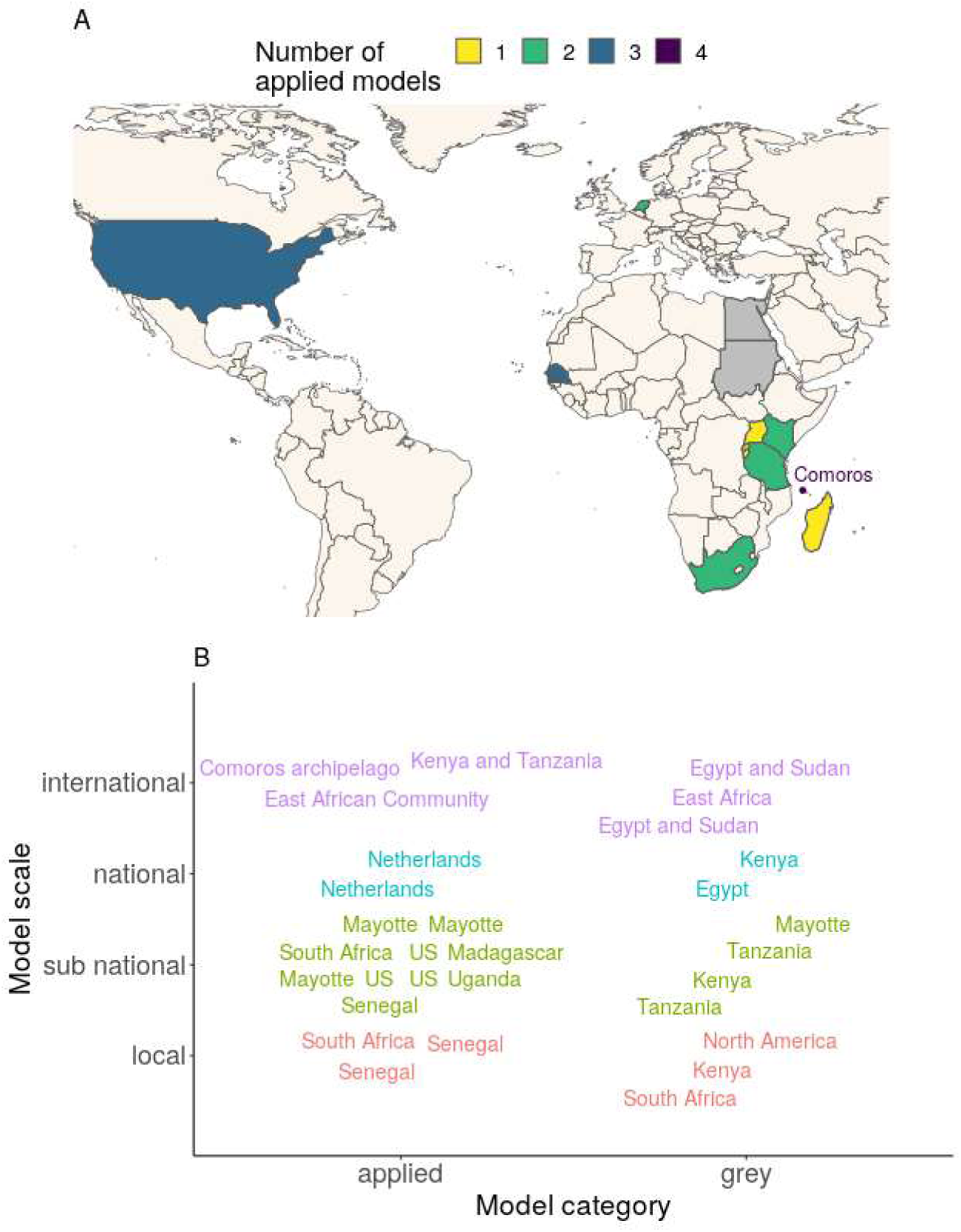
**A** - Location and number of RVF models. Grey models are mapped but not counted in totals because they sometimes refer to a non-precise context (e.g. East Africa, North America, see B). Locations of grey models which are not also studied in applied models are shown in grey (Egypt, Sudan). The point north of Madagascar, accompanied by text, is centered on the Comoros archipelago. It stands for 4 models applied to Mayotte island and 1 model applied to the whole Comoros archipelago, including Mayotte. **B** - Scale of applied and grey models. Labels represent model locations, with one label per model, hence sometimes repeated locations. Labels are colored to help identify the scale (y-axis). East African Community = Burundi, Kenya, Rwanda, and Tanzania. Besides, all 4 models applied to Mayotte considered the whole island (374 sq. km), but those models are classified as sub-national. Sub-figures A and B are not restricted to spatialized models

Spatial models, with at least two distinct locations, represented 45% (n = 22) of models (Table B in S1). Among those, 12 were applied, 5 theoretical, and 5 grey models (Table B in S1). All were discrete spatial models. Sixteen out of 22 spatial models incorporated connections between their spatial entities (Table B in S1): vertebrate hosts moved in 9 cases, vectors and hosts could move in 3 cases, and in 4 other cases, the connection was indirect, in the sense that the force of infection of one location was influenced by neighbors, taking into account distance, or prevalence. Three models were not spatialized but did include emigration and immigration of hosts (Table B in S1).

It should be noted that regions with recurring virus circulation, such as Mauritania, Mozambique, Namibia or Botswana [12] are still left out from the RVF modeling effort. Identifying the possible hurdles preventing model development in those regions is important. In addition, RVF being a transboundary animal disease, larger scale models are needed, able to gauge the role of animal movements in the transmission dynamics. Currently, international applied models do not incorporate connections between their spatial entities (Table B in S1), probably due to a lack of data. A coordinated data collection effort is required across affected countries, focusing on both commercial and pastoral mobility, and making these data easily accessible to epidemiological research teams.

#### Data

Data were used in 25 out of 49 (51%) models. Here, we define data as any raw information, as opposed to a readily available parameter value extracted from another study. Several types of data were used (Table C in S1): experimental (4/25), environmental (19/25), epidemiological (15/25), demographic (18/25), related to movements (6/25, Table B in S1), and geographical (6/25). Most (23/25) models used more than one type of data, and sometimes had several distinct datasets per type (Table C in S1). Among grey models, 7 used data and 6 did not.

We identified a total of 102 datasets (Table C in S1), corresponding to 4 datasets per model on average (102/25), ranging from 2 to 10. Only 44% of all datasets were available as spatialized data and used as such (with at least two distinct locations), and 45% as time-series (with at least two different time points) (Table C in S1). Regarding epidemiological datasets (n=24), 25% were spatialized, and 58% were time-series (Table C in S1). This is lower than environmental datasets (n=28), which were 57% spatialized and 86% time-series (Table C in S1). This supports conclusions made in recent reviews [16,17] which highlighted important gaps in RVF epidemiological data. Potential corrective measures would depend on whether such missing data is not collected or not made accessible. Most models with data managed to use at least one spatialized dataset (15/25, 60%) or time-series (23/25, 92%, Table C in S1). This indicates that mechanistic models can resort to all types of data to try and compensate for the lack of precision in epidemiological reporting. Five studies used epidemiological data not published elsewhere [37,41,47,48,61], showing that modeling studies can also be seen as a way to valorize new datasets.

We categorized data use into three categories: calibration, input, and model assessment. Calibration was defined as the parametrization of one process or initial condition of the model, transforming the data in some way. This was done in 17 cases (Table C in S1). Input was the fact of using the raw data directly as a parameter or initial condition of the model. This was done in 20 cases (Table C in S1). Model assessment referred either to parameter inference or qualitative estimation looking to maximize similarity between epidemiological model outputs and data. This was done in 12 cases (Table C in S1).

Ultimately, building accurate models helpful for policy makers requires the support of data. However, for RVF as well as for other infectious diseases, no single data source can be expected to inform each relevant parameter. Hence, the integration of information from many heterogeneous sources of data has become the norm [93]. This is a challenging task, as different datasets will be of different quality, potentially dependent, or in conflict [93]. Regarding quality, model-driven data collection can be a solution, but remains the exception rather than the rule [94]. Finally, we noted that in 39% of cases (4/10), models tailored to a location with known RVFV circulation, and which used epidemiological data, did not include any scientist from a local institution in their author list. This is important to highlight, at a time when concerns are being raised about the equity of South-North research collaborations [95,96].

#### Host and vector compartments

Most models (34/49) included a single vertebrate host, most of the time broadly labeled as livestock (25/34, Table 1). When two hosts were accounted for, it was most often done to add a human compartment (9/14, Table 1). Cecilia et al. (2020), Durand et al. (2020), Fischer et al. (2013) and Gachohi et al. (2016)[57,61,62,69] distinguished small ruminants (sheep and goats) from cattle. This grouping was made to incorporate differences in attractiveness to mosquitoes [57,61,62,69] or in RVF-induced mortality [57,61,69]. In addition to livestock, Barker et al. (2013) [43] included birds as incompetent hosts, used as alternate blood-feeding sources by vectors. The model by McMahon et al. (2014) [65] was the only one explicitly including a wildlife compartment, but did not describe the way the associated carrying capacity, (i.e. the maximum population size which can be sustained by the environment) was estimated based on land use data. Sumaye et al. (2019) [64] included a probability to pick up infection from wildlife hosts with a single parameter. Beechler et al. (2015) and Manore & Beechler (2015) [36,37] both modeled African buffaloes (*Syncerus caffer*), either captive or free-ranging.

The role of wildlife seemed largely understudied. Even if RVF virus circulation has been highlighted in several wildlife species, with clinical signs in some ruminants, the potential role of those species in the epidemiological sylvatic cycles in endemic areas is still poorly understood [97,98]. Studying the competence of local wildlife species for RVF virus transmission, along with their attractiveness to mosquitoes, is a prerequisite to determine the relevance of this question in a given territory [98–101].

In hosts, assumptions regarding the clinical expression of the disease varied. Chitnis et al. (2013), McMahon et al. (2014), Pedro et al. (2014) and Pedro et al. (2016) [39,45,65,77] included an asymptomatic state in hosts. Durand et al. (2020), Gachohi et al. (2016), Leedale et al. (2016), Taylor et al. (2016) and Tennant et al. (2021) [50,51,60,61,69] distributed hosts in age classes and (except Tennant et al. (2021) [60]) took into account differences in disease-induced mortality across classes. In Tennant et al. (2021) [60], only younger age classes moved between islands of the Comoros archipelago, and the initial proportion of immune individuals differed between classes. Cavalerie et al. (2015), Chamchod et al. (2014), Chamchod et al., (2016), Durand et al. (2020) and Sumaye et al. (2019) [41,61,64,70,78] incorporated abortion in livestock hosts due to RVF infection.

In terms of transmission routes, Cavalerie et al. (2015), Durand et al. (2020) and Nicolas et al. (2014) [41,59,61] included the possibility of direct transmission between vertebrate hosts. Among 11 models including a human compartment (Table 1), three [42,64,73] considered human-to-mosquito transmission, for a total of 10 models considering mosquito-to-human transmission, and one model [67] only considering livestock-to-human transmission. It would seem relevant for future models to evaluate the likelihood of humans being dead-end hosts, considering the difficulty to obtain direct observations on that matter.

Models with explicit vector compartments (43/49) included 1 (n = 20), 2 (n = 20) or more (n = 3) vector taxa (Table 1). Models with two taxa were all combining *Aedes* and *Culex* spp. vectors, while models with one taxon often did not precise the type of vector studied (10/20). The diversity of vector species was important, with the most common being *Ae. vexans* (n = 5) and *Cx. poicilipes* (n = 4). Pedro et al. (2017) [76] studied ticks (*Hyalomma truncatum*) in addition to *Aedes* and *Culex*. Sumaye et al. (2019) [64] included *Ae. mcintoshi, Ae. aegypti* and two generic *Culex* vectors in their model, distributed in different ecological zones of Tanzania. Cecilia et al. (2020) [57] included *Ae. vexans, Cx. poicilipes*, and *Cx. tritaeniorhynchus* distributed in different ecological zones of Senegal.

Eleven models (22%) incorporated the influence of abiotic factors on the life cycle and competence of vectors, with dedicated equations. Cecilia et al. (2020), EFSA AHAW Panel et al. (2020 – Model 2), Fischer et al. (2013), Leedale et al. (2016), Lo Iacono et al. (2018) and Mpeshe et al. (2014) [42,49 - Model 2,51,57,62,63] took into account the influence of temperature and/or rainfall on the lifespan of adult vectors lifespan. Gachohi et al. (2016), Leedale et al. (2016), Lo Iacono et al. (2018), Mpeshe et al. (2014), Xue et al. (2012) and Xue et al. (2013) [42,44,46,51,63,69] took into account the influence of temperature and/or rainfall on the egg laying rate and on the development or survival of aquatic stages. Barker et al. (2013), Cecilia et al. (2020), Fischer et al. (2013), Lo Iacono et al. (2018) and Mpeshe et al. (2014) [42,43,57,62,63] took into account the influence of temperature on the extrinsic incubation period (EIP) and on the biting rate (Durand et al. (2020) and EFSA AHAW Panel (2020 - Model 2) [49 - Model 2,61] on EIP only, Leedale et al. (2016) [51] on biting rate only). Further sophistications, including the dependence to water body surface, were included into Cecilia et al. (2020), Durand et al. (2020) and in Lo Iacono et al. (2018) [57,61,63]. For Cecilia et al. (2020) and Durand et al. (2020) [57,61], this was done indirectly by relying on an external entomological model for vector population dynamics [102]. Alternatively, in Métras et al. (2017), Métras et al. (2020) and Tennant et al. (2021) [47,48,60], the lack of data on vector densities urged the authors to use an environmental proxy (Normalized Difference Vegetation Index (NDVI) or rainfall) to drive vectorial transmission, without including an explicit vector compartment.

Dealing with multiple hosts and vectors makes it difficult to predict disease emergence, spread, and potential for establishment. It has been shown that accounting for a higher biodiversity in epidemiological models can result in amplification or dilution effects depending on species’ competence and abundance [103,104]. In the case of RVF, the role and contribution of hosts and vectors to transmission dynamics is largely understudied. Quantifying these roles is crucial to design targeted and efficient control strategies, and will require more knowledge on the intrinsic heterogeneity between host and vector species. Within-host modeling can help in this matter, but such models for RVF are rare [67,105]. Besides, a paramount hypothesis driving model behavior is the contact structure assumed between hosts and vectors, mathematically embodied by the force of infection.

#### Force of infection

We chose to focus on the diversity of functional forms (FFs) used in RVF models for the force of infection related to vector-borne transmission. This was applied only to models explicitly including a vector compartment. Among those, a majority (29/43) did not justify their choice of FF, even though the force of infection, as a disease transmission term, encapsulates authors’ assumption on the host-vector interactions, and therefore influences their predictions (Figure 5, [106]).

**Figure 5.**
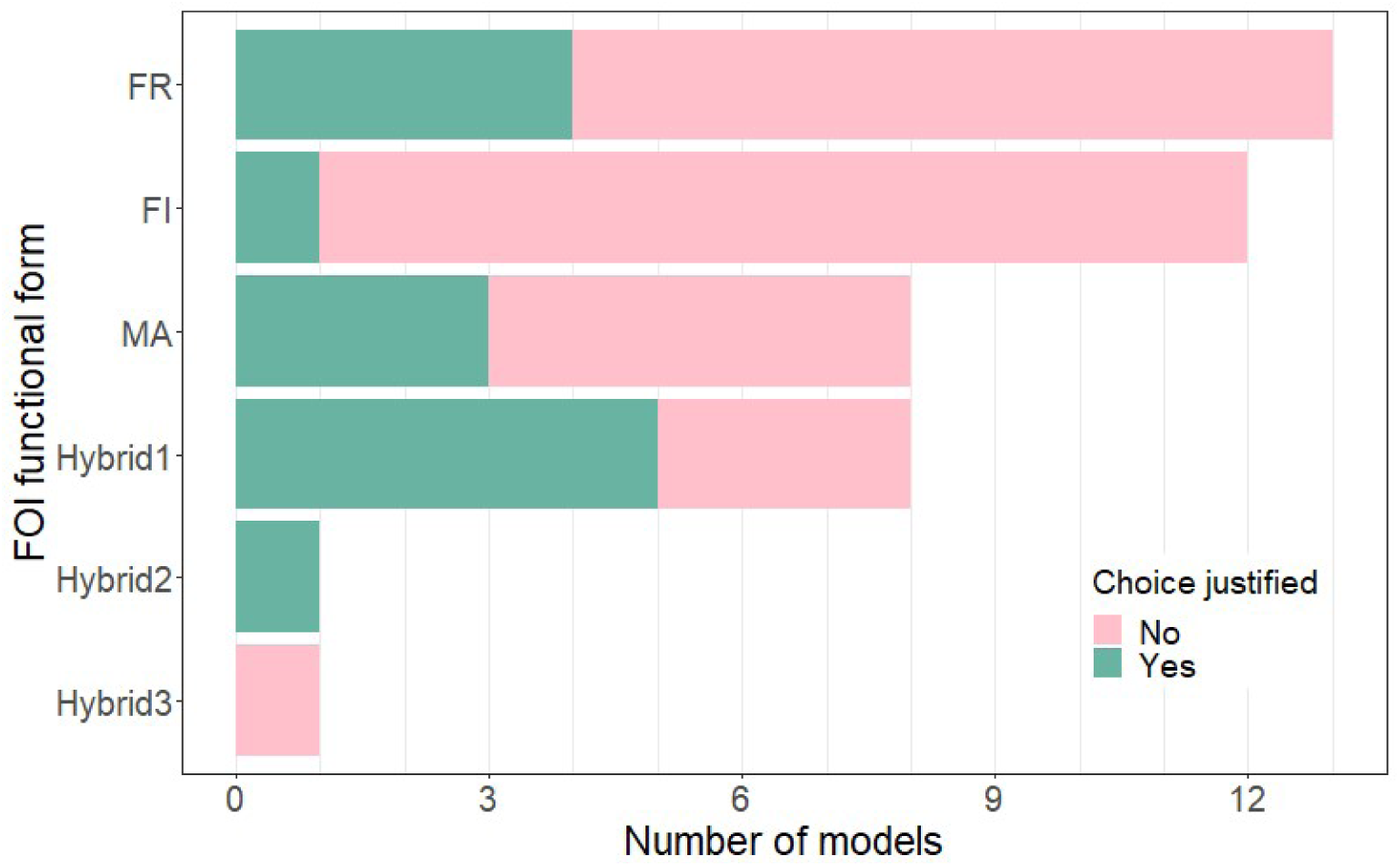
Functional forms (FFs) used by models for their force of infection (FOI; vector-borne transmission only). The full bar length indicates the number of models using a given FF, the color determines how many models properly justified their choice of FF. See Eq 1–6 for details on each FF. See Table 1 for details on papers using a given FF.

In standard SIR-type models, the force of infection (FOI) is the rate at which individuals go from the susceptible (S) state to the infectious (I, or exposed, E) state. Biologically, the FOI can be decomposed as *p*_contact_. *p*_inf_. *p*_tran_. For vector-borne transmission, *p*_contact_ is the contact rate between vectors (subscript *v*) and hosts (subscript *h*), *p*_inf_is the probability that a given contact is with an infectious individual, and *p*_transm_ is the probability that a contact with an infectious individual results in successful transmission. This can be declined in two directions of transmission: vector-to-host and host-to-vector, which affects the value of these parameters. For *p*_inf_, we have:

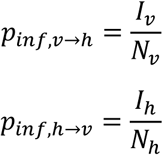

The value of *p*_transm_ can also vary depending on the source and target of the infection, but is not linked to host nor vector densities, but rather individual-level parameters (e.g., species, viremia, immune response). The different functional forms which can be seen in vector-borne disease models then arise from different assumptions on *p*_contact_ [107]. Six FFs were found in reviewed models (Table 1, Figure 5). We detail them in Eq. 1–6.

### Reservoir frequency-dependence

The reservoir frequency-dependent (FR, n = 13, Eq. 1) functional form assumes that the rate at which a vector bites hosts is constant across host (reservoir) densities (i.e. the vector does not feed more if there are more hosts), while the number of bites received by a host is proportional to the current vector-to-host ratio (i.e. a host is fed upon more if surrounded by more mosquitoes, at constant host population). Consequently, we get:

- *p*_contact,h→v_ = *a*, with a being the biting rate, usually defined as the maximal rate allowed by the gonotrophic cycle (i.e the minimum time required between blood meals for a female to produce and lay eggs). This results in 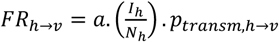.
- 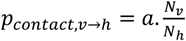 which simplifies with *p*_inf,h→v_ and results in 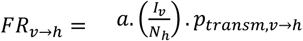.

We can write, using *β*_hv_ and *β*_vh_ as aggregated terms, sometimes called adequate contact rates as in Gaff et al. (2007) [32]:

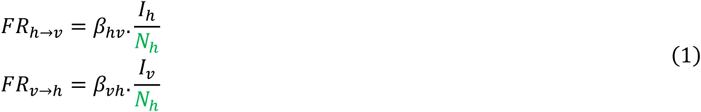

This functional form (FF) is therefore called reservoir frequency-dependent because the total number of hosts is on the denominator for both transmission directions (*v* → *h* and *h* → *v*). With this FF, the vector-to-host transmission rate linearly increases with the vector-to-host ratio, and can therefore reach unrealistic values. Indeed, at some point, hosts are expected to deploy defense mechanisms to protect themselves from biting, preventing the vector population from getting all the blood meals needed.

### Mass action

The mass action functional form (MA, n = 8, Eq. 2), sometimes called pseudo mass action, is density-dependent. It assumes that a vector bites hosts at a rate proportional to the number of hosts, and that a host is bitten at a rate proportional to the number of vectors. Consequently, we get:

- *p*_contact,h→v_ ∝ *N*_h_, ignoring a possible constant derived from the previous biting rate a, which simplifies with *p*_inf,h→v_ and results in *MA*_h→v_ ∝ *I*_h_. *p*_transm,h→v_.
- *p*_contact,v→h_ ∝ *N*_v_, which similarly gives *MA*_v→h_ ∝ *I*_v_. *p*_transm,v→h_.

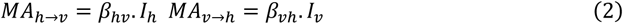

With this functional form, the biting rate of vectors per unit time can exceed their physiological capacity above certain host densities, which again, becomes unrealistic.

### Infected frequency-dependence

Following the nomenclature by Wonham et al. (2006) [106], who presented susceptible frequency dependence, we describe the infectious frequency-dependent (FI, n = 12, Eq. 3) functional form. It assumes that the rate at which a vector bites hosts is constant across host (reservoir) densities, while the number of bites received by a host is constant across vector densities. The transmission terms are then both correlated to the proportion of infectious in the population:

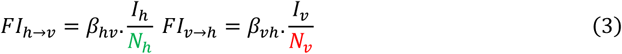

The only plausible situation inducing a constant contact rate in both directions is one where the vector-to-host ratio remains constant. Indeed, if we assume that a vector systematically gets the blood meals it physiologically needs, and modeled hosts are the sole source of blood, then a shortage in hosts (high vector-to-host ratio) should result in an increase in bites per host (FR functional form). Alternatively, if the number of bites received per host is saturated (and constant) to account for their defense mechanism, then a vector’s biting rate should vary with host densities, depending on whether this constrained system allows it to feed as it needs.

### Alternative functional forms

Functional forms FR, MA, and FI can only apply biologically at certain population densities, outside which they can generate aberrant values and therefore lead to erroneous predictions [106]. Three alternative FFs were found in RVF models to prevent this issue (Figure 5). Those FFs require additional parameters to constrain the contact rate between populations.

The first alternative FF (Eq. 4, Hybrid1 in Table 1 and Figure 5, n = 8) was first used in RVF models by Chitnis et al. (2013) [39], who previously formulated it in a model of malaria transmission [108].

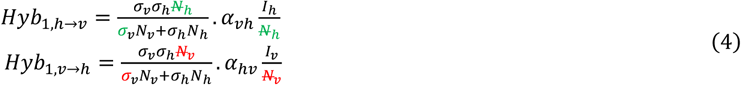

In Eq. 4, *α*_hv_ and *α*_vh_ refer to probabilities of successful transmission given contact, from host to vector and vice versa. *σ*_v_ is defined as the maximum number of times a mosquito would bite a host per unit time, if freely available. This is a function of the mosquito’s gonotrophic cycle and its preference for a given host species. *σ*_h_ is the parameter added to avoid abnormally high contact rates and represents the maximum number of bites sustained by a host per unit time. Although *σ*_h_ seems virtually impossible to estimate in the field, this alternative FOI can efficiently prevent erroneous model predictions and has therefore often been reused in RVF models. It is also the most justified FF (5/8, Figure 5). Some slight variations in its mathematical formulation can be found in Sumaye et al. (2019) [64].

The second alternative FF was used by McMahon et al. (2014) [65] (Eq. 5, Hybrid2 in Table 1 and Figure 5, n = 1).

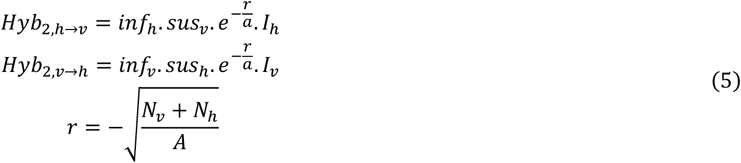

Here, *inf* and *sus* refer to a vector or host infectivity and susceptibility, respectively. The contact rate is formulated as 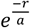, with *a* the characteristic length of local spread. In *r, A* is the patch area.

A last alternative FF was used in Lo Iacono et al. (2018) [63] (Eq. 6, Hybrid3 in Table 1 and Figure 5, n = 1).

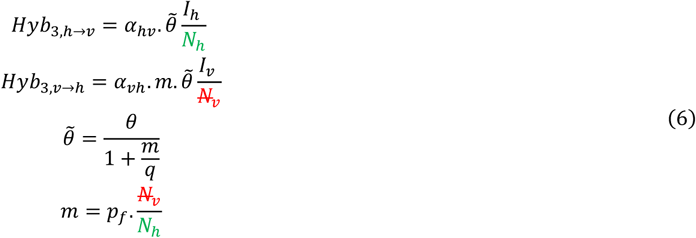

Here, *p*_*f*_ is the proportion of the mosquito population able to detect and feed on the host species under consideration and *m* is therefore an “effective” vector-to-host ratio. ^∼^*θ* is the biting rate, function of *θ*the rate of completion of the gonotrophic cycle, *m*, and of *q*, the vector-to-host ratio for which vector fecundity is divided by 2, to account for the decrease in fecundity in the case of absence of sufficient hosts to take a blood meal.

In 82% of cases (14/17), a model used the same functional form (FF) for its force of infection as its parent model (Figure 2). In 9/14 cases, the parent model did not justify the choice of FF used, and no further justification was provided in the subsequent model in 7/9 cases. In 3/17 cases, the FF was changed compared to the parent model, which induced a justification in 2/3 cases. In addition, in 3 cases, the representation of vectors was implicit in a model and its parent model, therefore preventing the classification of the force of infection into any FF.

Several review papers on various epidemiological models concluded that the choice of a functional form for the force of infection could greatly affect model behavior. Begon et al. (2002), Hoch et al. (2018) and McCallum et al. (2001) [107,109,110] focused on non-vectorial transmission. Hopkins et al. (2020) [111] focused on parasite transmission, which could be through a vector, but did not include possible variations in frequency-dependent functions. Wonham et al. (2006) [106] focused on FFs used to model vectorial transmission of West Nile virus and also noticed an important diversity. In 2001, McCallum et al. were already recommending to “explicitly state and justify the form of transmission used” as well as “evaluate several alternative models of transmission, if possible” [110]. Contact structures between host and vector populations are hard to observe in natural conditions. This should be an additional incentive for modelers to explicitly state the reasoning behind their choice of functional form, which can be motivated by the context of their case study (e.g., expected host and vector densities, mixing between the populations). A comparison of the FF listed presently would be useful. The conclusions might vary depending on whether this is done through theoretical scenarios, keeping all other parameters equal, or by fitting different models to a common empirical dataset. The latter might not be able to discriminate between FF to select the best-performing one, because of underlying correlations between parameters.

## Conclusion

In the last five years, more mechanistic models of RVF virus transmission dynamics have been published (n=26) than in the ten previous years combined (n=23). This possibly indicates a growing interest for RVF epidemiology, although it is known that the number of publications is continuously growing in all fields [112,113]. This could also reflect an increasing trust in mechanistic models in the field of infectious disease epidemiology (Lofgren et al., 2014; Ezanno et al., 2020). Our review highlighted important knowledge gaps, rarely addressed in mechanistic models of RVF transmission dynamics. In our opinion, the most pressing issues are i) the incorporation of heterogeneity among host and vector species, in order to determine their relative role in transmission dynamics, which will require a focus at the within-host scale, and ii) the development of large scale models, able to quantify the role of animal mobility in RVF spread. Both of these research avenues will rely on novel data sets being generated, and will require methodological accuracy and transparency, particularly with regards to the choice of force of infection.

Importantly, we note that only 6 studies made their code available (Table 1), which represents 19% of models published since 2015. Adopting this practice more broadly would increase the reproducibility of results and encourage the community to bring existing work further.

## Data Availability

All relevant data are within the manuscript and its Supporting Information.

## Supporting information

**S1 Supporting information - Reading grid and complementary tables**. Text A: Reading grid; Table A: Vaccination strategies implemented in models and main results; Table B: Characteristics of spatial models as well as non-spatial models with external renewal; Table C: Type of datasets and their use.

## Text A: Reading grid

### Research questions, context, main analyses and results

Main modeling objective (primary/secondary possible): understand; anticipate; control Model category: theoretical; applied; grey

Geographical location: if applicable, precise if zone with history of RVF or RVF-free

Context under study: hypothetical incursion or real world setting; endemic or epidemic scenario; other

Geographical scale: international; national; sub-national; local

Is a parent model clearly stated?

What is common/different from the parent model?

Indicators of disease spread: R (any type of reproductive ratio, precise if analytical, from data, by simulation); number of individuals in a given health state; summary statistic; other

Model outputs: spatial patterns; temporal patterns; sensitivity of the model to parameters/hypothesis; parameter estimation; other

Control measures tested Model selection performed?

Sensitivity analysis performed? If yes, which type: one-at-a-time; global Main results

Limits of the model (as presented in the paper)

### Data

If presence of data,

Type of data: experimental; environmental; epidemiological; clinical; demographic; movement; other

If the data spatialized, is it used as such in the model?

If the data presented as time-series, is it used as such in the model?

Use of data: calibration; model selection; input; inference/model assessment

Origin of data: all published or open access; some are published for the first time in peer review

### Hosts

How many vertebrate taxa were considered? What taxa?

What health states?

Is there population renewal?

Were age class considered?

If yes, what type of age classes?

Is the herd/human population detailed further (male/female; pregnant; occupation; other)? Important differences between host taxa or host types within a taxa : population density/dynamics; movement patterns; attractiveness to mosquitoes; infectiousness/susceptibility; clinical outcome; duration of latent/infectious period; development of immunity; some were dead-end hosts; other Clinical outcomes described : asymptomatic/symptomatic; mild/severe; disease-induced mortality; abortion; none/other

### Vectors

How was the vector component modeled: implicitly (no state variable); explicitly (at least one state variable)?

How many taxa?

What taxa?

Main characteristics of the vector population and differences between taxa, precise when driven by abiotic factor: life cycle (emergence, feeding behavior); infection (probability, extrinsic incubation period); transmission (probability, vertical transmission)

What health states?

How was the aquatic population modeled: not at all; implicitly (emergence rate of adults described by a parameter or function); explicitly (at least one state variable); other (e.g. extracted from an external model)?

If implicit aquatic population, type of parameter/function: constant; forced (e.g. sinusoidal, precise if informed by data); input-dependent (e.g. abiotic variable); other

If explicit aquatic population, features (which state, influence of abiotic factors, possible dormancy etc.)

Blood meal frequency: constant; variable (precise with what factor); not a parameter

Blood meal distribution among hosts: homogeneous; heterogeneous; not a parameter

Is there an explicit factor besides host taxa driving heterogeneous biting (host size, behavior, infection status etc.)?

### Spatiotemporal dynamics and modeling paradigm

How many spatial locations: one with no immigration or emigration/location was undefined or vaguely defined (spaceless model); more than one location or model included terms describing immigration or emigration?

Type of spatial model: discrete number of locations/pixels; continuous space; connectivity network among individual hosts; other

Resolution (surface covered by one pixel, if applicable) What moves: nothing; vertebrate hosts; vectors; both?

If hosts move, what type of movement: commercial; nomadic; other?

If vectors move, what type of movement: follow hosts; random; other (e.g. wind)? Duration of simulations, and if known, time period (month, year)

Timestep

Characteristics of the model: deterministic/stochastic; agent-based/compartmental If stochastic: what is stochastic, number of runs etc.

### Transmission and infection

Functional form of the force of infection (FOI, for vector-host transmission) Is the choice of functional form justified/discussed?

Transmission parameter: aggregated, constant; aggregated with variation (function with no clear biological translation, e.g. proxy for seasonality); decomposed (different parameters for biological processes such as biting rate, probability of successful transmission, feeding preferences etc.) Other routes of transmission (precise FOI if applicable): mosquito-mosquito; livestock-livestock; mosquito-human (1 way or 2 ways); livestock-human (1 way)

Hypothesis for time spent in E/I/R state, for hosts/vectors: exponential; other (precise if driven by abiotic factor)

**Table A:**
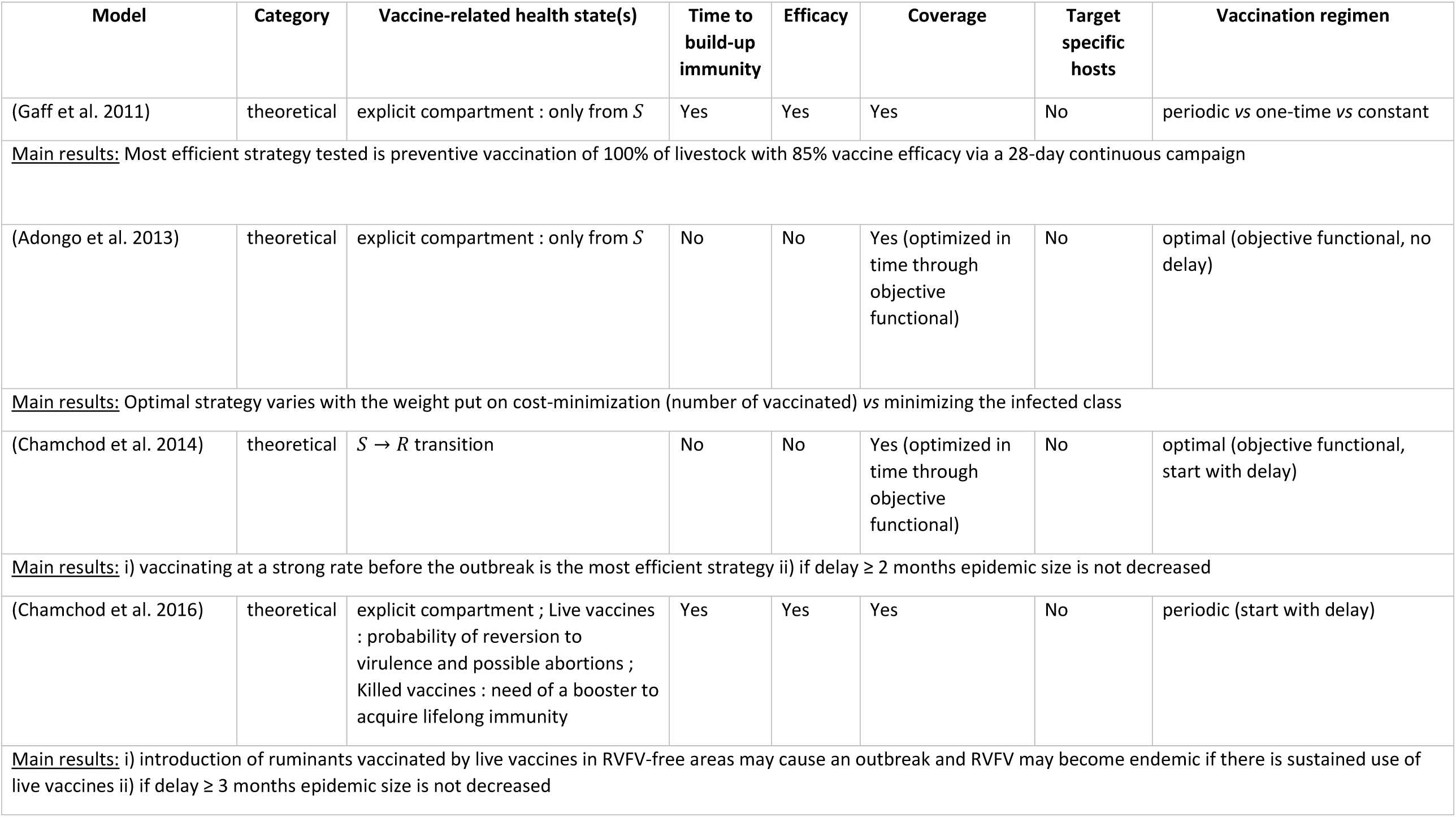
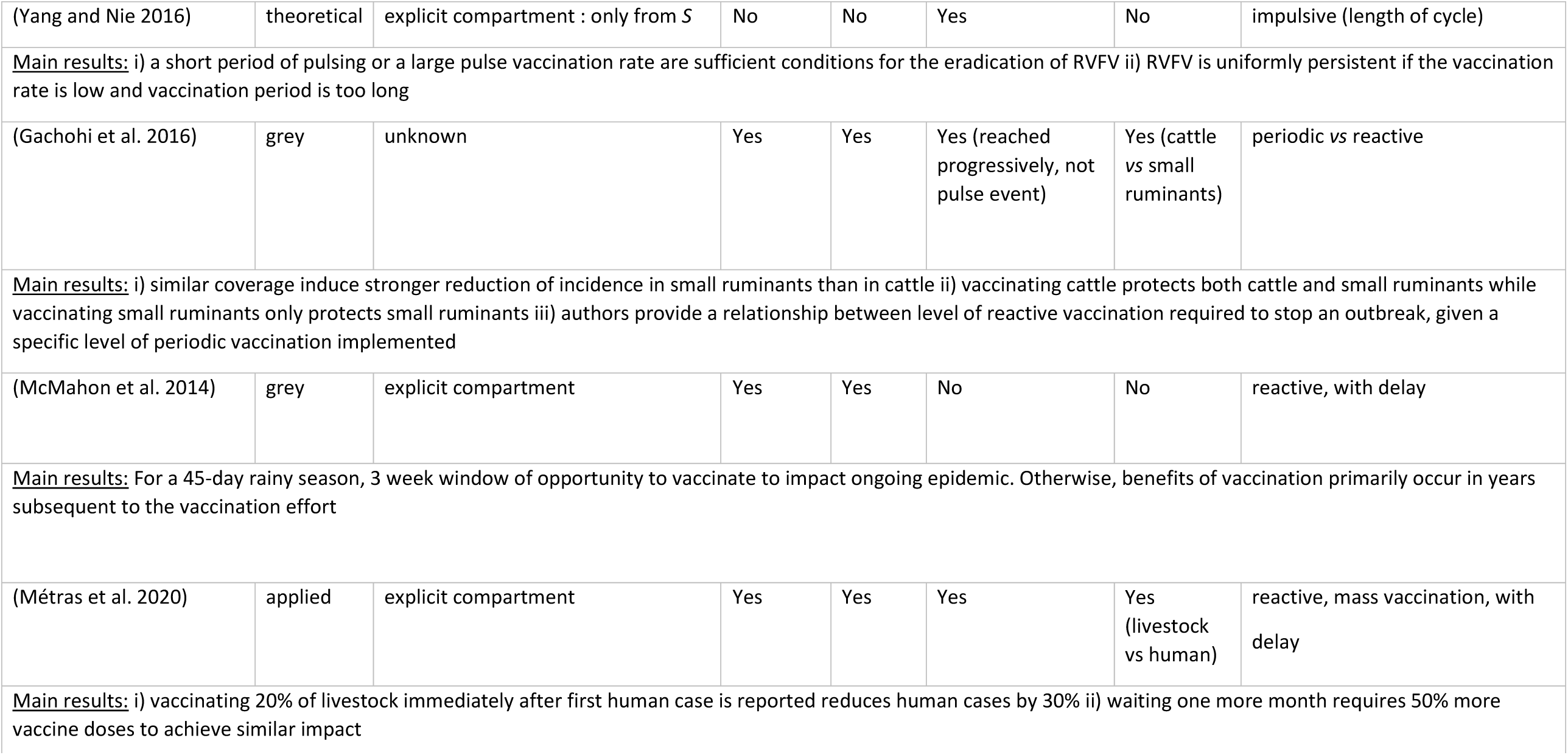
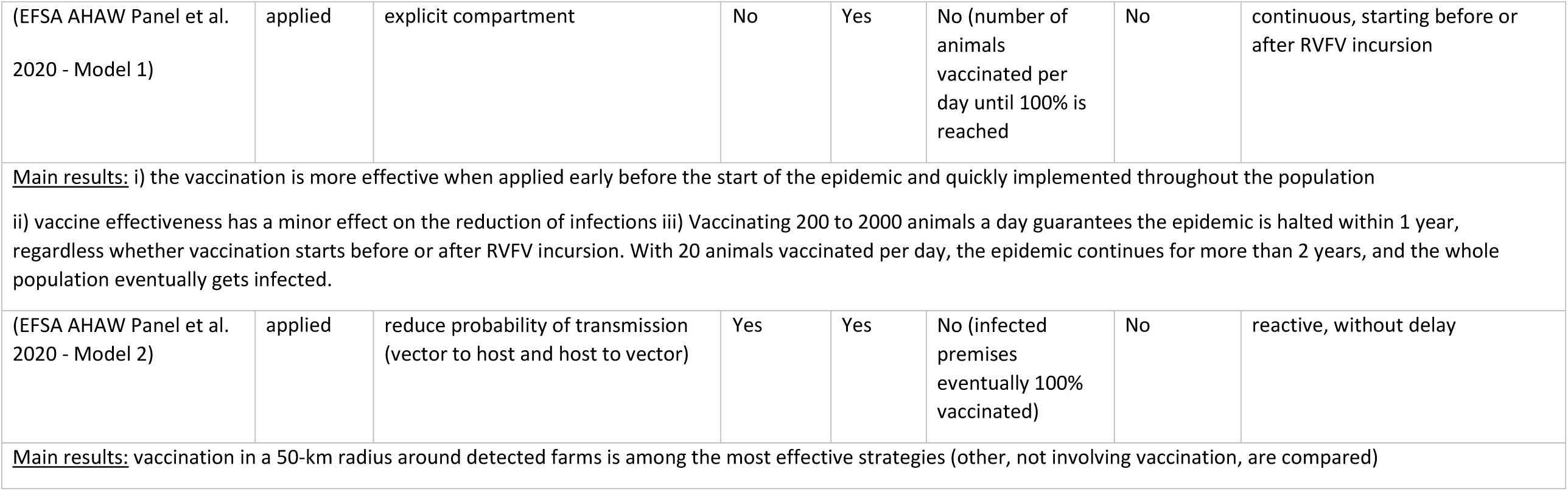
Vaccination strategies implemented in models and main results.

**Table B:** Characteristics of spatial models as well as non-spatial models with external renewal. Without any geographical context, we chose not to assign a scale to theoretical models.

| Reference | Category | Location | Scale | Connection | Movement data / use of proxy |
| --- | --- | --- | --- | --- | --- |
| **Spatial models** |  |  |  |  |  |
| (Niu et al. 2012) | theoretical |  |  | vectors and hosts move | None |
| (Xue and Scoglio 2013) | theoretical |  |  | hosts move | None |
| (Xue and Scoglio 2015) | theoretical |  |  | indirect through FOI | None |
| (Wen, Teng, and Liu 2019) | theoretical |  |  | hosts move | None |
| (Python Ndekou Tandong et al. 2020) | theoretical |  |  | hosts move | None |
| (Gao et al. 2013) | grey | Egypt and Sudan | international | hosts move | None |
| (McMahon et al. 2014) | grey | East Africa | international | indirect through FOI | None |
| (Mpeshe, Luboobi, and Nkansah-Gyekye 2014) | grey | Tanzania | sub-national | disconnected entities | None |
| (Xiao et al. 2015) | grey | Egypt and Sudan | international | hosts move | None |
| (Sumaye et al. 2019) | grey | Tanzania | sub-national | hosts move | None |
| (Xue et al. 2012) | applied | South Africa | sub-national | vectors and hosts move | vectors : wind<br>livestock : animals sold, number in feedlots<br>humans : distance, population, commuting and return rates |
| (Xue and Scoglio 2013) | applied | US | sub-national | vectors and hosts move | maximum percentage of animals moving, extensive agricultural setting |
| (Barker et al. 2013) | applied | US | sub-national | disconnected entities | None |
| (Fischer et al. 2013) | applied | Netherlands | national | disconnected entities | None |
| (Nicolas et al. 2014) | applied | Madagascar | sub-national | hosts move | renewal practices of cattle breeders : trade, barter, number of animals, origin |
| (Leedale et al. 2016) | applied | Kenya and Tanzania | international | disconnected entities | None |
| (Taylor et al. 2016) | applied | East African Community | international | disconnected entities | None |
| (Scoglio et al. 2016) | applied | US | sub-national | hosts move | None |
| (Sekamatte et al. 2019) | applied | Uganda | sub-national | indirect through FOI | None |
| (Cecilia et al. 2020) | applied | Senegal | sub-national | disconnected entities | None |
| (EFSA AHAW Panel et al. 2020 - Model 2) | applied | Netherlands | national | indirect through FOI | None |
| (Tennant et al. 2021) | applied | Comoros archipelago (including Mayotte) | international or national : to be determined | hosts move | expert knowledge of local veterinary services to inform prior distribution of movement parameters (start date, duration, trade estimates, infectious imports) |
| **Non spatial, open models** |  |  |  |  |  |
| (Gil et al. 2016) | grey | Egypt | national | hosts entry (importations) | None |
| (Métras et al. 2017) | applied | Mayotte | sub-national | hosts entry | unofficial trade |
| (Durand et al. 2020) | applied | Senegal | local | hosts in and out | survey of nomadic herders : main stops, dates |

**Table C:** Type of datasets and their use. A number is assigned to each dataset in column “Type of data” and used in subsequent columns for readability.

| Study | Type of data used (numbers defined in this column are used in the following columns) | Spatialized data | Time series data | Use of data |  |  |
| --- | --- | --- | --- | --- | --- | --- |
|  |  |  |  | Calibration | Input | Inference / model assessment |
| (Métras et al. 2017) | **Environmental**<br>- Vegetation (1)<br>**Epidemiological**<br>- Serological (livestock) (2)<br>**Demographic**<br>- Livestock + age structure (3)<br>**Movement**<br>- Importation (4)<br>- Starting date of import + duration (5) |  | 1, 2 | 3, 4 | 1 | 2, 5 |
| (Métras et al. 2020) | **Environmental**<br>- Rainfall (1)<br>**Epidemiological**<br>- Serological (livestock) (2)<br>- Serological (human) (3)<br>- Incidence (human) (4) |  | 1, 4 | 2,3 | 1 | 2, 4 |
| (Cecilia et al. 2020) | **Environmental**<br>- Temperature (indirect) (1)<br>**Demographic**<br>- Livestock (2)<br>- Vectors (3)<br>**Other**<br>- Vector trophic preferences (4) | 1, 2, 3 | 1, 3 | 4 | 1, 2, 3 |  |
| (Durand et al. 2020) | **Environmental**<br>- Temperature (indirect) (1)<br>**Epidemiological**<br>- Serological (livestock, age structured) (2)<br>**Demographic**<br>- Livestock (3)<br>- Vectors (4)<br>**Movement**<br>- Nomadic movements (5) |  | 1, 2, 4 | 3, 4, 5 | 1, 4 | 2 |
| (Leedale et al. 2016) | **Environmental**<br>- Temperature (1)<br>- Rainfall (2) | 1, 2, 3, 4, 6 | 1, 2, 3, 4, 6 |  | 1, 2, 5 | 3, 4, 6 |
|  | **Epidemiological**<br>- Outbreaks dates (3)<br>- Mosquito surveillance (infection) (4)<br>**Demographic**<br>- Livestock (5)<br>- Vectors (6) |  |  |  |  |  |
| (Taylor et al. 2016) | **Environmental**<br>- Temperature (1)<br>- Rainfall (2)<br>**Other**<br>- Social vulnerability (3) | 1, 2, 3 | 1, 2 |  | 1, 2, 3 |  |
| (McMahon et al. 2014) | **Environmental**<br>- Land use (1)<br>- Rainfall (2)<br>**Demographic**<br>- Livestock (3)<br>- Humans (4) | 1, 2, 3, 4 | 2 | 1, 2 | 3, 4 |  |
| (Mpeshe, Luboobi, and Nkansah-Gyekye 2014) | **Experimental**<br>- Temperature dependent survival rates (1)<br>**Environmental**<br>- Temperature (2)<br>- Rainfall (3) | 2, 3 | 2, 3 | 1 | 2, 3 |  |
| (Fischer et al. 2013) | **Environmental**<br>- Temperature (1)<br>**Demographic**<br>- Livestock (2)<br>- Vectors (abundance) (3)<br>- Vectors (dynamics) (4) | 2, 3 | 1, 4 | 4 | 1, 2, 3 |  |
| (Nicolas et al. 2014) | **Environmental**<br>- Land use (1)<br>**Epidemiological**<br>- Serological (livestock) (2)<br>**Demographic**<br>- Livestock (density) (3)<br>- Livestock (calving and mortality) (4)<br>- Vectors (5)<br>**Movement**<br>- Trade and barter (6)<br>**Geographical**<br>- Villages locations (7) | 1, 2, 3, 6, 7 | 5 | 4, 5 | 1, 3, 6 | 2 |
| (Barker et al. 2013) | **Experimental**<br>- EIP duration variations with temperature (1)<br>**Environmental**<br>- Temperature (2)<br>- Land use (3)<br>**Demographic**<br>- Livestock (4)<br>- Vectors (5)<br>- Birds (6) | 2, 3, 4, 6 | 2, 5 | 1, 5 | 2, 3, 4, 6 |  |
| (Gachohi et al. 2016) | **Environmental**<br>- Rainfall (1)<br>**Epidemiological**<br>- Vaccination data (2)<br>- Outbreaks dates (3)<br>**Demographic**<br>- Livestock + age structure (4) | 1 | 1 | 1, 2, 3, 4 | 1 |  |
| (Pedro, Abelman, and Tonnang 2016) | **Epidemiological**<br>- Prevalence (livestock) (1)<br>- Outbreaks years (2) |  | 1, 2 |  |  | 1, 2 |
| (Cavalerie et al. 2015) | **Epidemiological**<br>- Outbreaks start (1)<br>- Duration of presence of the virus (2)<br>- Serological (animals) (3)<br>**Demographic**<br>- Vectors (4) |  | 3 | 1, 4 |  | 2, 3 |
| (Bicout and Sabatier 2004) | **Environmental**<br>- Rainfall (1)<br>**Epidemiological**<br>- Rate of decay of IgG positive individuals (livestock) (2)<br>**Demographic**<br>- Vectors (3) |  | 1, 3 | 1, 2, 3 |  |  |
| (Tuncer et al. 2016) | **Experimental**<br>- TCID50, IgM, IgG in sheep (1)<br>**Epidemiological**<br>- Incidence (humans) (2) |  | 1, 2 |  |  | 1, 2 |
| (Scoglio et al. 2016) | **Demographic**<br>- Livestock (1)<br>- Vectors (2)<br>**Geographical**<br>- Farms locations (3) | 1, 3 |  | 2 | 1, 3 |  |
| (Xue et al. 2012) | **Environmental**<br>- Temperature (1)<br>- Rainfall (2)<br>- Wind (3)<br>**Epidemiological**<br>- Incidence (livestock) (4)<br>- Incidence (humans) (5)<br>**Demographic**<br>- Livestock (6)<br>- Human (7)<br>**Movement**<br>- Trade (8)<br>- Commuting/return rates (humans) (9)<br>**Geographical**<br>- Distance between patch centers (10) | 3, 4, 5, 10 | (1, 2 : assumed),<br>4, 5 |  | 1, 2, 3, 6, 7, 8, 9, 10 | 4, 5 |
| (Xue et al. 2013) | **Environmental**<br>- Temperature (1)<br>- Rainfall (2)<br>**Demographic**<br>- Livestock (3)<br>**Movement**<br>- Aggregated movements (livestock) (4)<br>**Geographical**<br>- Distance between node centers (5) | 3, 5 | 1, 2 | 4 | 1, 2, 3, 5 |  |
| (Beechler et al. 2015) | **Epidemiological**<br>- Serological (animals) (1)<br>- BTB status (2) |  | 1 |  |  | 1, 2 |
| (Sekamatte et al. 2019) | **Demographic**<br>- Livestock (1)<br>**Geographical**<br>- Centroids (longitude and latitude) (2) | 1, 2 |  |  | 1, 2 |  |
| (EFSA AHAW Panel et al. 2020 - Model 1) | **Environmental**<br>- Rainfall (1)<br>**Epidemiological**<br>- Serological (livestock) (2)<br>**Demographic**<br>- Livestock (3) |  | 1, 2 | 2 | 1, 3 |  |
| (EFSA AHAW Panel et al. 2020 - Model 2) | **Environmental**<br>- Temperature (1)<br>**Demographic**<br>- Livestock (2) | 1, 2, 3 | 1 |  | 1, 2, 3 |  |
|  | **Geographical**<br>- Farms locations (3) |  |  |  |  |  |
| (Tennant et al. 2021) | **Environmental**<br>- Vegetation (1)<br>**Epidemiological**<br>- Serological (livestock) (2)<br>**Demographic**<br>- Livestock (3)<br>**Movement**<br>- Trade (4) | 1, 2, 3 | 1, 2 | 4 | 1, 3 | 2 |
| (Lo lacono et al. 2018) | **Experimental**<br>- Larvae and pupae mortality rate according to temperature (1)<br>**Environmental**<br>- Temperature (2)<br>- Water body surface (3)<br>**Epidemiological**<br>- Duration of interepidemic periods (4) |  | 2, 3 | 1 | 2, 3 | 4 |

